# Convergent coexpression reveals shared biological mechanisms underlying common and rare variant risk in six neuropsychiatric disorders

**DOI:** 10.1101/2025.08.13.25333593

**Authors:** Hanna Abe, Calwing Liao, Lide Han, Theodore Morley, Michael E. Talkowski, Kristen J. Brennand, Douglas M. Ruderfer

**Author notes:** Correspondence &.

## Abstract

Genome-wide association studies (GWAS) and large-scale rare variant burden analyses have identified both common and rare loss-of-function variants associated with neuropsychiatric disorders. Yet, the shared biological processes influenced by both classes of variation remain poorly characterized. In this study, we utilized transcriptomic data from 933 post-mortem brain samples to identify genes that show convergent coexpression with GWAS and rare variant burden risk genes across six neuropsychiatric disorders. Despite largely distinct sets of significant risk genes from GWAS and rare variant burden studies, we found a significant overlap in their convergently coexpressed genes. These convergent genes showed enrichment for common and rare variant heritability and highlighted key biological pathways and cell-type markers impacted by both types of genetic variation. Compared to genes coexpressed with one variant class, shared convergent genes exhibited stronger evolutionary constraint and greater enrichment for known drug targets, underscoring their potential therapeutic relevance. Collectively, our results establish a systematic and generalizable framework for integrating coexpression data with genetic risk to reveal transcriptional programs supported by both common and rare variant evidence, offering mechanistic insights into neuropsychiatric diseases.

## Introduction

Understanding how genetic variation contributes to disease remains a fundamental challenge in human genetics. Numerous common and rare genetic variants have been discovered for complex diseases, yet the underlying biological mechanisms remain largely elusive^1,2,3^. Integrating evidence from both genome-wide association studies (GWAS) and whole exome sequencing (WES) has enhanced the interpretation of disease-associated variants and aided in identification of high confidence genes for therapeutic development and functional investigation^1,4,5,6,7^. Rare variant data has also been valuable to prioritize candidate genes within GWAS loci that may play a causal role in disease^2,4^. Pathways influenced by both common and rare variants likely reflect core disease mechanisms, biological processes essential to disease onset and progression, with relevance across genetically heterogenous patient populations^8,9^.

Efforts to uncover shared functional mechanisms between common and rare variants remain limited by methodological challenges and insufficient statistical power of current GWAS or rare variant burden studies^10,11^. Other factors such as extreme trait polygenicity and bias toward longer genes in rare variant burden studies make systematic investigation of shared mechanisms influenced by both common and rare variants challenging^10,11^. Beyond assessing overlap of gene level associations, integrating transcriptomic changes driven by common and rare genetic variants in disease relevant tissues can illuminate their shared functional impact on disease-relevant pathways. Accordingly, we propose that a systems-level approach, incorporating transcriptomic effects of GWAS and rare variant burden-associated genes, may uncover fundamental biological processes underlying disease.

A primary strategy to capture downstream transcriptomic effects of risk genes involves experimentally perturbing the genes through methods such as CRISPR. This is typically followed by RNA-sequencing to identify genes with altered transcriptional profiles resulting from the perturbation. Several studies have successfully identified downstream differentially expressed genes following the perturbation of select disease risk genes^2,12^. However, scaling this approach to hundreds of genes implicated from GWAS and rare variant burden studies remains both technically challenging and cost prohibitive. To address this, recent studies have explored computational or *in silico* approaches to infer downstream transcriptional effects. One practical alternative is using gene coexpression in disease relevant tissues. Within a given tissue, two genes are coexpressed if they exhibit significantly correlated gene expression levels across samples. Disease associated genes often exhibit such coordinated expression, making coexpression networks a valuable tool for uncovering their functional relationships^13,14,15^. Liao et al showed that the coexpression profile of a gene significantly correlated with differential expression level from a CRISPR experiment. Furthermore, meta-analysis of coexpression profiles across multiple disease genes, an approach known as convergent coexpression, has been shown to capture the shared downstream effects of different gene perturbations and implicate novel risk genes^16^.

Previous studies have shown that rare and common variants associated with a given disease could impact similar gene networks resulting in the system wide dysregulation of downstream genes. This is evident in Gandal *et al* and Walker *et al*, where common and rare variants associated with ASD were enriched in similar coexpression modules^13,17^. Identifying the network level regulation could highlight disease relevant processes and pathways. Convergent coexpression of common and rare variant risk genes allows for the identification of genes that are consistently coexpressed with both variant types in a context specific manner, providing a well-powered and systematic framework to explore their shared biological mechanisms.

In this study, we sought to identify the shared biological mechanisms between GWAS and rare variant burden risk genes across six brain disorders. Leveraging post-mortem brain transcriptomic data, we identified genes that were convergently coexpressed with both GWAS and rare variant burden risk genes, thereby extending our analysis beyond genome-wide significant loci. We evaluated the disease relevance of these convergent coexpression genes, demonstrating their contribution to both SNP-based and burden heritability. Notably, genes convergent with both GWAS and burden gene sets were under stronger evolutionary constraint and were enriched for known drug targets. Furthermore, we uncovered shared biological pathways between using genes convergently coexpressed with GWAS and rare variant burden risk genes, revealing insights not captured by genomic data alone and highlighting common processes across brain disorders.

## Methods and materials

### Data sources and collection

We sourced publicly available summary statistics for GWAS and rare variant burden studies using exome data for six brain phenotypes: Alzheimer’s (AD)^18,19^, autism spectrum disorder (ASD)^3,20^, bipolar disorder (BD)^21,22^, epilepsy (EPI)^23,24^, Parkinson’s disease (PD)^25,26^ and schizophrenia (SCZ)^27,28^ from Psychiatric Genomics Consortium (PGC), the GWAS catalog, or study specific database. A full list of the studies and corresponding summary statistics used here can be found in Table 1.

**Table 1.**
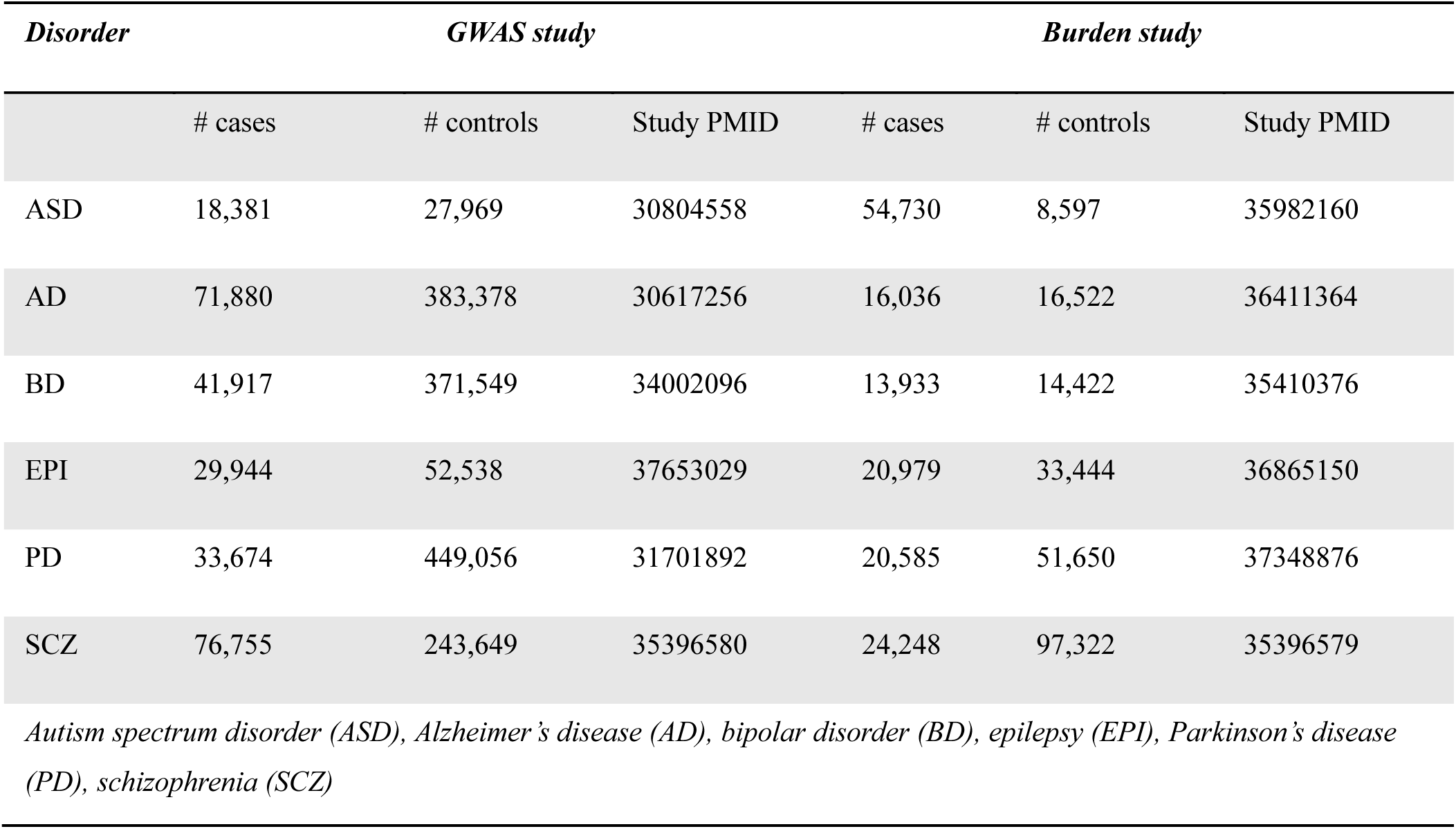
GWAS and rare variant burden summary data used in this study.

| Disorder | GWAS study |  |  | Burden study |  |  |
| --- | --- | --- | --- | --- | --- | --- |
|  | # cases | # controls | Study PMID | # cases | # controls | Study PMID |
| ASD | 18,381 | 27,969 | 30804558 | 54,730 | 8,597 | 35982160 |
| AD | 71,880 | 383,378 | 30617256 | 16,036 | 16,522 | 36411364 |
| BD | 41,917 | 371,549 | 34002096 | 13,933 | 14,422 | 35410376 |
| EPI | 29,944 | 52,538 | 37653029 | 20,979 | 33,444 | 36865150 |
| PD | 33,674 | 449,056 | 31701892 | 20,585 | 51,650 | 37348876 |
| SCZ | 76,755 | 243,649 | 35396580 | 24,248 | 97,322 | 35396579 |

Autism spectrum disorder (ASD), Alzheimer’s disease (AD), bipolar disorder (BD), epilepsy (EPI), Parkinson’s disease (PD), schizophrenia (SCZ)

### Gene prioritization from GWAS and rare variant burden studies

Next, we identified risk genes from the collected GWAS and rare variant burden summary statistics for the six brain disorders. Since rare variant burden studies include gene level association results, we identified genes that passed the exome-wide significant threshold. Depending on the total number of genes in the study, the threshold after multiple testing correction ranged from pvalue < 2.2e-6 to p < 1.7 e-3. Genes that passed the study specific significance threshold were considered rare variant burden risk genes.

To prioritize risk genes from the GWAS summary statistics, we applied four widely adopted gene annotation approaches; nearest gene, MAGMA^29^ (Multi-marker Analysis of GenoMic Annotation), PoPS^30^ (Polygenic Priority Score), and TWAS^31^ (Transcriptome Wide Association Study). To identify the nearest gene for each lead independent SNP, we first collected the LD independent SNPs from supplementary files of each of the corresponding GWAS. We then determined the transcription start site (TSS) based on the strand orientation using GENCODE^32^ version 44 for GRCh37 or hg38. Next, we computed the distance from each SNP to genes on the same chromosome and designated the gene with the shortest distance as the nearest gene. We also identified the second nearest gene using the same approach for comparison. To run MAGMA on GWAS summary statistics we first performed SNP-to-gene mapping by considering SNPs located 1 kb upstream of the TSS and 0.5 kb downstream of the transcription end site (TES). This mapping was based on SNP positions from the 1000 Genomes European reference panel^33^ and gene locations from hg19 or hg38 depending on the genome build of summary statistics. Gene-based p-values were then calculated. For TWAS, we utilized PrediXcan models trained on GTEx brain tissues. Using the S-prediXcan^34^ approach, we applied logistic regression to test the association of the GWAS summary statistics with 12 brain PrediXcan models applying Bonferroni correction. To prioritize genes using PoPS, we first assessed gene-level associations with MAGMA, leveraging LD estimates from the 1000 Genomes European data. Next, we conducted enrichment analysis for gene features listed in the PoPS Github page (https://github.com/FinucaneLab/gene_features). Finally, PoPS scores were computed by fitting a joint model that accounts for the enrichment of all identified features. Genes with the highest PoPS score for each lead SNP were chosen. We further evaluated concordance of genes prioritized across the five approaches. For each of the six brain phenotypes, we used Jaccard similarity index to compare similarity of genes identified. The Jaccard index, defined as the size of the intersection divided by the size of the union of two sets, provides a measure of similarity ranging from 0 (no overlap) to 1 (complete overlap).

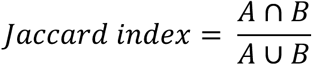

where, A and B are sets of genes identified across any two gene prioritization approaches. ∣A∩B∣ is the number of genes common to both sets (the intersection) and ∣A∪B∣ is the total unique number of genes in either set (the union).

### Convergent coexpression analysis

To assess convergent coexpression among the prioritized genes from GWAS or rare variant burden studies, we utilized transcriptome data from the CommonMind Consortium (CMC), which included RNA-sequencing data from 933 postmortem brain samples from dorsolateral prefrontal cortex (DLPFC). RNA-sequencing data was processed following methods described in Han et al. (2020)^35^ and Liao et al. (2023)^16^. Pairwise gene expression correlations were computed using Pearson’s method for 17,062 genes in the CMC dataset, with correlation coefficients transformed into Z-scores via Fisher transformation. To quantify convergent coexpression, we applied Stouffer’s weighted Z-score method to meta-analyze coexpression Z-scores across GWAS or rare variant burden risk genes for each of the six phenotypes. Genes in the top 10% of absolute convergent coexpression Z-score were considered as convergently coexpressed or convergent genes. We applied similar processing steps to assess convergent coexpression using transcriptomic data from GTEx v8 tissues^36^.

### Heritability analysis for coexpression convergent genes

S-LDSC (Stratified Linkage Disequilibrium SCore regression)^37^ was used to estimate the level of heritability (h^2^) explained by common variants in convergent genes. This method was developed to partition polygenic trait heritability by one or more functional annotations. Common variants (MAF ≥ 0.05) were annotated for a set of genes within a convergent coexpression decile bin, using a default genomic window of 1 kb that extends 500 bp on either side of each gene. S-LDSC was applied to annotated SNPs to compute population-specific LD scores and quantify the complex trait heritability captured by our convergent gene annotations. Standardized effect size (*τ*\*) was used to evaluate how well the annotations capture variation. This metric is comparable across annotations and is defined by the proportionate change in per-SNP *h*^2^ associated with a one standard deviation increase in the value of the annotation. The effect size of a particular annotation is evaluated controlling for the effects of other annotations. The approach quantifies effects that are unique to a given annotation. The effect size for the annotation with greater true causal variant membership will be larger and more significantly positive. A value of *τ*\* greater than zero suggests that membership to the annotation increases per-SNP h^2^.

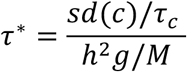

where, *h^2^g* is the heritability due to all SNPs in baseline, *M* is the total number of snps in the regression model, *sd(c)* -- standard deviation increases in annotation *c* and *τ* is the coefficient output from heritability. Significance (P value) of standardized effect size is quantified by assuming *τc*\**/se* (*τc*\*) follows a normal distribution with mean 0 and variance of 1 (*τ*\**/se* (*τc* *) *∼ N* (*0*, 1) as done previously in Kim *et al*, 2019^38^, Gupta *et al*,2023^39^, and Ravichandran *et al*,2024^40^.

We also investigated enrichment in heritability explained by rare variants within convergent genes from both GWAS and rare variant burden studies. Burden heritability regression (BHR) R package v0.1.0 was used to estimate the phenotypic variance explained by gene-wise burden of rare coding variants^41^. We used the default settings and baseline model (LD v2.2) provided at the BHR Github page (https://github.com/ajaynadig/bhr). The BHR performed regression of the burden test statistic on the burden score using summary statistics of the association analysis and allele frequencies at the variant level and derived the burden heritability through estimation of the regression slope.

### Enrichment of evolutionarily constrained and functionally prioritized gene-sets

To assess the biological relevance of convergent genes, we examined their overlap with several gene sets associated with functional importance. These included genes intolerant to loss-of-function mutations (high pLI scores), genes known to be haploinsufficient, genes targeted by FDA-approved drugs, and genes implicated in recessive disorders obtained from previously published sources^38,40^. Excess overlap was performed as implemented in Kim *et al*, and Ravichandran *et al*, to estimate enrichment of convergent coexpression genes^38,40^. To measure excess overlap, we define as follows:

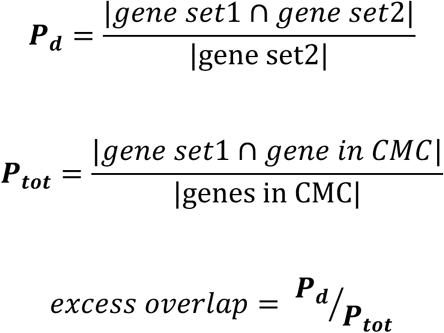

where geneset1 is the list of genes in the essential gene category and gene set2 is the list of genes in the coexpression convergent decile bin. We also collected known drug targets for the six disorders from Minikel et al^42^ and Open Targets database^43^and performed excess overlap with convergent genes.

### Functional annotation of convergent genes

Gene Ontology (GO) analysis of genesets that are in 90^th^ percentile or above (top decile) of convergent coexpression Zscore for both GWAS and rare variant burden disease genes for all six brain disorders were performed using cluster-Profiler R package v.4.8.2 (GO)^44^. Biological processes that have Benjamini-Hochberg (BH) adjusted pvalue < 0.05 for both GWAS and rare variant burden convergent genesets were considered shared processes. Cell type enrichment analysis was conducted on shared convergent genes, defined as those in the top decile of convergence Z-scores from both GWAS and rare variant burden analyses. Enrichment was performed using the Enrichr platform, with default settings from the Azimuth_Cell_Types_2021 and Cell_Marker_2024 databases^45^. Results with an FDR-adjusted p-value below 0.05 were considered statistically significant.

### Cross-disorder comparison of convergent genes and processes

To assess whether convergent genes of a given brain disorder are associated with the other five disorders, we conducted Fisher’s exact enrichment analysis on discovered genes associated with the six brain disorders. The analysis was performed using the top convergent genes, identified based on the absolute values of GWAS and rare variant burden convergence Z-scores. To investigate whether enriched biological processes supported by both GWAS and rare variant burden convergence for one disorder are also shared across other disorders, we performed GO enrichment analysis using genes convergent in both GWAS and rare variant burden or each of the six disorders independently using clusterProfiler. We then extracted processes that were identified in both gene categories specifically. These processes were then aggregated into clusters based on semantic similarity using the R package rrvgo (v1.12.21). For each resulting cluster, we calculated the median enrichment rank of processes in that cluster to summarize significance. Visualization was performed using the R package pheatmap (v1.0.12), and each cluster was labeled with a representative “Parent Term” based on the hierarchy of the GO terms, summarizing its functional theme.

## Results

### Limited overlap of GWAS and rare variant burden risk genes in six brain disorders

We compiled recent large-scale summary statistics from studies of six neuropsychiatric disorders: Autism spectrum disorder (ASD), Alzheimer’s disease (AD), bipolar disorder (BD), epilepsy (EPI), Parkinson’s disease (PD), schizophrenia (SCZ) (Table 1). The six neuropsychiatric disorders were selected based on having at least five LD-independent index SNPs or genes surpassing the significance threshold in both GWAS and rare variant burden analyses, respectively.

For rare variant burden studies, genes carrying loss-of-function rare variants that surpassed the exome-wide significant threshold were identified (Table S1). For GWAS studies, we applied four widely used methods to link variants to genes: nearest gene, polygenic priority score (PoPS)^30^, MAGMA^29^, and TWAS^31^ (Figure 1A, Table S2). We used pairwise Jaccard similarity coefficients to evaluate how consistently the four approaches implicated the same genes and found limited overlap of implicated genes across the approaches (Figure 1B, Figure S1). The highest overlap was observed between TWAS and MAGMA, followed by PoPS and nearest gene approach, with average Jaccard index of 0.15 and 0.1 respectively. The lowest overlap was observed between PoPS and TWAS with average index of 0.01 (Figure 1B). GWAS gene prioritization methods also differed in their level of overlap with rare variant burden genes. The nearest gene approach showed the strongest overlap, with significant enrichment observed in AD (Fisher’s exact p = 1.2 × 10^−7^), SCZ (p = 0.01), and PD (p = 0.001). PoPS and MAGMA also showed significant overlap, with PoPS yielding enrichment in AD (p = 4.2 ×10^−5^) and SCZ (p = 0.009), and MAGMA in AD (pvalue = 6.6 × 10^−7^) and EPI (pvalue = 0.003). In contrast, TWAS approach showed no overlap with rare variant burden genes in any disorder (Figure 1C).

**Figure 1.**
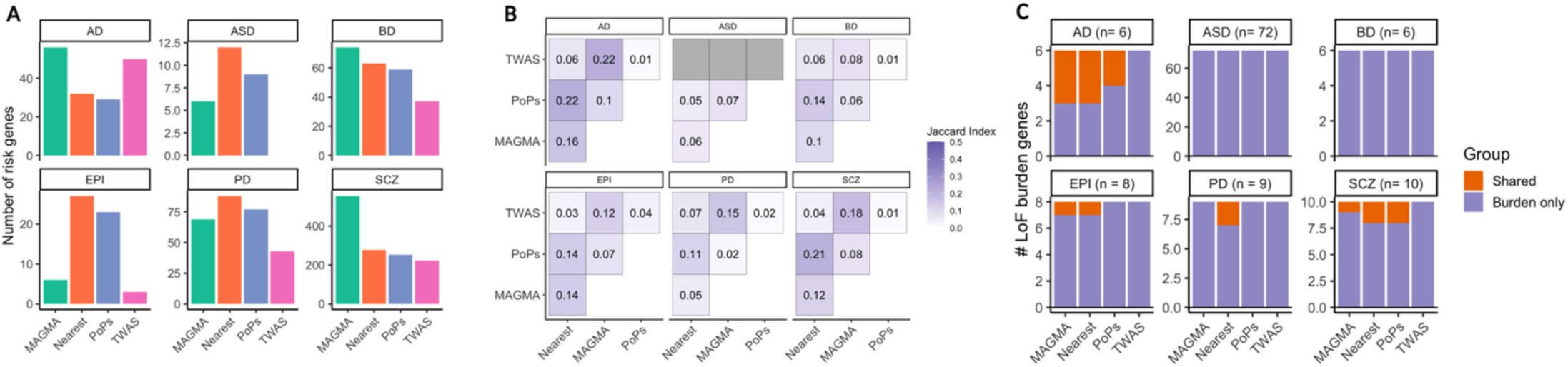
Genes prioritized from rare variant burden and GWAS studies across six brain disorders. A) Number of prioritized genes from GWAS across four approaches in six brain disorders B) Pairwise similarity comparison of GWAS-prioritized genes across approaches C) Counts of rare variant burden genes, highlighting overlaps with GWAS-prioritized genes across approaches

### Convergent coexpression of GWAS risk genes captures disease specific patterns

In our previous work (Liao et al.^16^), we demonstrated convergent coexpression for rare variant burden risk genes of ASD and SCZ using transcriptome data from prefrontal cortex tissue collected from the CommonMind Consortium (CMC). Here, we extended the analysis to rare burden studies of four additional brain disorders and updated the ASD geneset using the latest data with 72 risk genes^3^. We quantified significance of overall convergence by comparing the variance of the distribution of convergent coexpression to variance of permuted random genes. Four of the six disorders, ASD, BD, EPI, and SCZ showed significant (empirical p < 0.05) overall convergence. Convergence level varied considerably across disorders. ASD showed the highest level of convergence (mean absolute convergence Z-scores = 1.02) followed by SCZ (0.74), EPI (0.67), BD(0.44), AD(0.285), and PD(0.221) (Table S3 & Figure S4). Convergence analysis in 12 brain tissues from GTEx revealed strong concordance with CMC results (median correlation = 0.75), particularly in cortical regions and the hippocampus (Figures S2 and S4).

Convergent coexpression analysis was then extended to the GWAS-prioritized genes across six brain disorders. Given the methodological differences among gene prioritization approaches, we evaluated convergence separately for each method and included the second-nearest gene as a negative control. PoPS showed the highest mean absolute convergence (0.642), followed by the nearest gene (0.452), MAGMA (0.420), second-nearest gene approach (0.298), and TWAS (0.262) (Figure S3). Based on these results, PoPS was selected for downstream analyses. GWAS level convergence also showed variation across disorders, with SCZ and BD exhibiting the highest overall convergence (SCZ_var = 4.39, BD_var = 0.95) while ASD showed the lowest (0.11) (Table S4 and Figure S4).

In light of limited overlap between GWAS and rare variant burden risk genes, we next sought to investigate the degree of overlap using convergently coexpressed genes. Applying Fisher’s exact test across the top deciles (>90^th^ percentile Zscore) of convergent genes, we observed significant overlap between the top GWAS and rare variant burden convergent genes across all six brain disorders. SCZ showed the highest degree of overlap (log2_Fisher’s OR = 6.64, p < 2.3 × 10^−269^), whereas PD showed the weakest (log2_Fisher’s OR = 2.76, p = 3.15 × 10^−196^) (Figure S8A). We also found that correlations between GWAS and burden convergence Z-scores were tissue dependent showing significantly stronger relationships in brain tissues than in non-brain tissues (median ρ_brain = 0.83, median ρ_non-brain = 0.40; Wilcoxon p-value = 2 × 10^−5^; Figure S8B).

### Convergent genes are enriched for both SNP-based and burden heritability

To assess enrichment of heritability among convergently coexpressed genes, we ranked genes into deciles based on their absolute convergent coexpression Z-scores. Using Stratified LD Score Regression (S-LDSC) (with baseline LD v2.2), we estimated the contribution of each geneset to SNP heritability, quantifying the standardized effect size (τ*). Genes in the highest convergence deciles showed the greatest enrichment for SNP heritability compared to other decile bins (τ* value range: 0.02 - 0.2) (Figure 2A, Table S5). We also measured heritability due to rare variants (burden heritability) using “BHR” approach. Similar to the common variant heritability, the most convergent genes with GWAS or rare variant burden risk genes showed the greatest enrichment for burden heritability (bhr heritability range: 0.00017 - 0.007) (Figure 2B). Genes with positive convergence (convergence Z-score > 0) showed greater bhr enrichment (enrichment; ASD = 2.05, EPI = 4.1) compared to negatively convergent genes (Z-score < 0) (bhr enrichment; ASD = 0.87, EPI = 0.51) (Figure S7, Table S6). Top convergent genes also showed high excess overlap with genesets from loss-of-function intolerant PLI) genes (mean excess overlap = 1.568, SE = 0.061), haploinsufficient (mean excess overlap = 1.64, SE = 0.33), and FDA approved drug target genes (mean excess overlap = 1.42, SE= 0.232) (Figure 2C, Figure S5, and Table S7).

**Figure 2.**
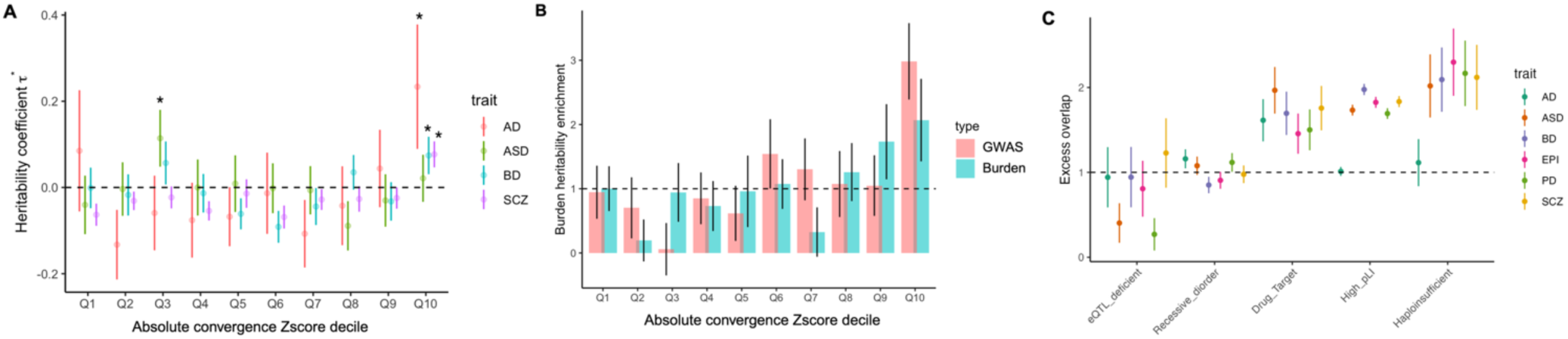
Enrichment of convergently coexpressed genes in disease heritability and curated genesets. A) Standardized heritability coefficient (*τ*\*) computed using s-LDSC for genesets grouped by absolute convergent coexpression signature with genes prioritized from PoPS B) Burden heritability quantifying enrichment for genes grouped in deciles of GWAS and rare variant burden absolute convergence Zscore in BD C) Overrepresentation of various gene sets in the top 10% of genes with the highest absolute convergence Z-scores.

### Shared convergent genes exhibit greater intolerance and drug target enrichment

We characterized genes that were uniquely convergent in either GWAS or rare variant burden analyses, as well as those convergently coexpressed with both sets of genes, referred to as “shared convergent genes”. Shared convergent genes showed greater intolerance, indicated by lower LOEUF scores (median LOEUF = 0.54), than genes uniquely convergent with GWAS (0.681) and rare burden (0.653) risk genes, with significant differences between shared and uniquely convergent genes (Wilcoxon p = 7.5 × 10^−10^) but not between GWAS and rare burden convergent genes (p = 0.3) (Figure 3A, Figure S8). Since LOEUF scores have been associated with gene length, we also explored other evolutionary constraint scores such as gismo^46^, Shet^47^, and pHaplo^48^. Consistent with above results, shared convergent genes were significantly more constrained compared to uniquely convergent genes in either GWAS (Wilcoxon p: gismo = 4 × 10^−3^, Shet = 2.1 × 10^−10^, pHaplo = 7.6 × 10^−5^) or rare variant burden analyses (Wilcoxon p: gismo = 1 × 10^−7^, Shet = 1.4 × 10^−10^ and pHaplo = 1.8 ×10^−3^) (Figure S8C).

**Figure 3.**
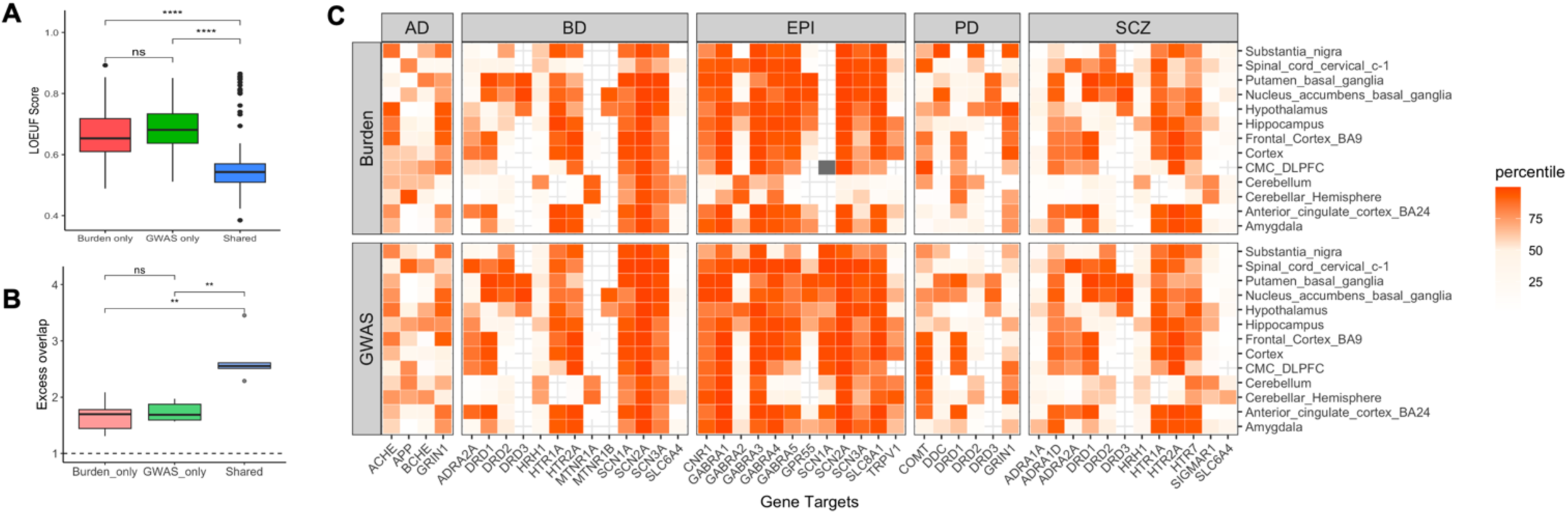
Intolerance level and drug target enrichment of convergently coexpressed genes. A) Median LOEUF score for uniquely and shared convergent genes (top decile convergence Z-score) B) Excess overlap of known drug target genes across all clinical phases (1-IV) in convergent genes C) GWAS and burden convergence level (percentile rank) of “launched” drug targets collected from Minikel et al^42^

To evaluate the therapeutic potential of shared convergent genes, we curated known drug target genes linked to the six brain disorders from the Open Targets database and Minikel et al., 2024^42,43^. For each disorder, we assessed the enrichment of known drug targets among genes exhibiting GWAS, burden, or shared convergence. Using targets across all clinical phases, we found that shared convergent genes exhibited stronger enrichment for known drug targets (mean excess overlap = 2.92) compared to genes uniquely convergent with either GWAS (1.74) or burden (1.66) risk genes (Figure 3B, Figure S9). To test whether this enrichment persists across different stages (phases) of drug development, we conducted a similar analysis after stratifying gene targets by the highest clinical phase reached. Shared convergent genes primarily showed significantly higher enrichment for disease-matched drug targets (mean excess overlap = 2.9, Wilcoxon p = 0.01) compared to those of other diseases (mean excess overlap = 1.66).

Since evolutionary constraint scores are commonly used to prioritize drug targets, we tested whether convergent coexpression offers additional explanatory power. We fitted two logistic regression models to assess the relationship between constraint and drug target status, before and after including convergent coexpression. Incorporating both GWAS and burden convergence significantly improved the model’s ability to explain drug target status compared to using constraint alone (ANOVA p-value range: 1.64 × 10⁻² to 1.73 × 10⁻¹³) (Table S8). Moreover, approved (“Launched”) drug targets showed similar level of convergence with both GWAS and rare risk genes in CMC and GTEx brain datasets. Drug targets, like ***SCN2A***, showed consistent convergence across brain regions, while other targets, such as ***DRD2*** and ***DRD3***, displayed region-specific convergence, highlighting the tissue specific nature of convergence (Figure 3C).

### Convergent coexpression uncovers shared processes between GWAS and rare variant burden risk genes

Convergent genesets enabled us to systematically test biological processes associated with both GWAS and rare variant burden risk. GO enrichment analysis on top convergent genes for each disorder revealed distinct and overlapping processes between the two types of genetic risk (Figure 4). In AD, *positive regulation of angiogenesis* (p = 1.8 × 10^−8^) and *extrinsic apoptotic signaling* (p = 8.42 × 10^−8^) were enriched exclusively in GWAS convergent genes while *endocytosis* (p = 1.1 × 10^−6^) and *axonogenesis* (p = 4.12 × 10^−9^) were uniquely enriched in burden-convergent genes. Notably, immune-related processes like *myeloid leukocyte activation and differentiation* (p_GWAS_ = 3.43 × 10^−16^, p_burden_ = 5.72 × 10^−8^) were enriched in both GWAS and burden convergent genes. Processes related to *synaptic vesicle transport*, *dendritic function*, and *neuronal development* were significantly enriched (adjusted pvalue < 0.05) among both GWAS and rare variant burden convergent genes in ASD, BD, EPI, PD, and SCZ (Table S9). *Neuronal migration processes* (p = 1.77 × 10^−7^) were uniquely enriched in burden convergent genes in ASD, whereas *cardiac muscle and heart growth* (p = 6.21 × 10^−5^) were specifically enriched in GWAS convergent genes for EPI. Additionally, burden convergent genes in PD showed enrichment for pathways related to *NFkB regulation and apoptotic signaling* (p = 2.51 × 10^−3^).

**Figure 4.**
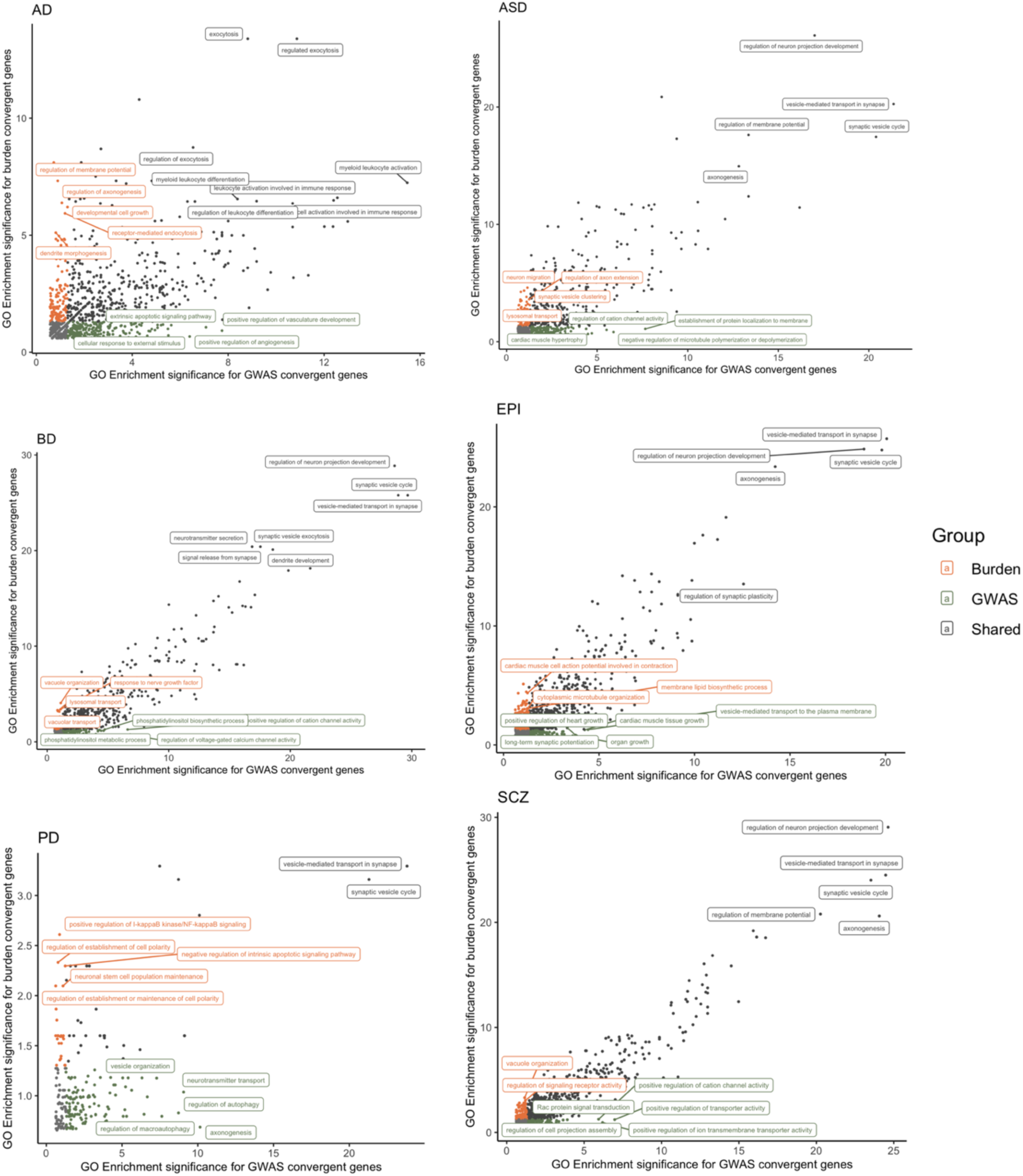
Shared biological processes enriched among GWAS and rare variant burden convergent genes. Colors indicate whether enrichment is specific to GWAS-convergent genes, burden-convergent genes, or shared between both. The x and y axis show -log10(enrichment pvalue).

### Convergent genes reveal overlapping cell types and biological processes across disorders

Shared convergent genes also showed overlapping cell type enrichment across the six brain disorders. Astrocyte marker genes were consistently enriched across all six disorders (ASD_p = 2 × 10^−5^, AD_pval = 0.013, BD_p = 4.69 × 10^−6^, EPI_p = 1.3 × 10^−6^, PD_p = 3.5 × 10^−9^, SCZ_p = 4.6 × 10^−6^) (Figure 5A). In contrast, microglia (AD_p = 7.2 × 10^−171^) and macrophages (AD_p = 1.07 ×10^−19^) markers showed disease-specific enrichment, primarily in AD. Glutamatergic neuron markers were enriched in the shared convergent genes for ASD (p = 0.02), BD (p = 0.01), EPI (p = 3 × 10^−4^), and SCZ (p = 6.3 × 10^−4^). Those four brain disorders also showed notable enrichment in Oligodendrocyte progenitor cell markers (ASD_pval = 8 × 10^−5^, BD_p = 4.7 × 10^−6^, EPI_p = 1.32 × 10^−7^, and SCZ_p =6.9 × 10^−8^) (Table S10).

**Figure 5.**
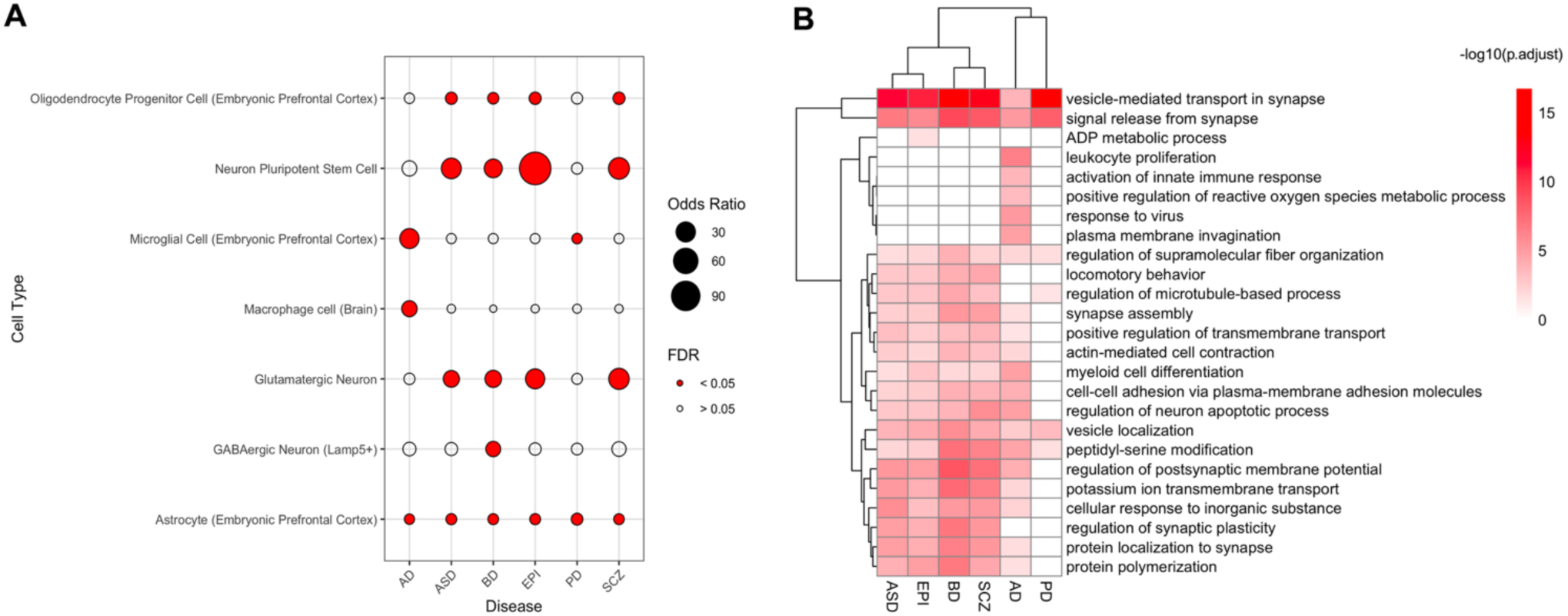
Cross disease comparison of convergent genes and biological processes. A) Over-representation analysis of cell type marker genes within the shared convergent gene sets B) Clustering of biological processes jointly supported by GWAS and rare variant burden convergence. Each cluster is labeled by the parent biological process term and shows the median enrichment significance across disorders.

In our results above, we had identified biological processes enriched in both GWAS and burden convergent genes for each trait separately. Next, we sought to assess which of the biological processes were shared across the six disorders. To reduce dimensionality, we performed semantic similarity on all biological processes that showed enrichment in both GWAS and rare variant burden convergent gene level using rrvgo R package. Some processes showed enrichment in only certain disorders such as *leukocyte* proliferation (median p = 1 × 10^−9^) and *innate immune system activation* (median p = 1.72 × 10^−7^) in AD. However, we observed a large cluster of processes involving *synaptic vesicle transport, signal release from synapse, vesicle localization,* and *peptidyl-serine modification* enriched in all six disorders (Figure 5B).

We examined how GWAS and burden convergent genes relate to known disease-associated genes, both within and across diseases. Genes prioritized from the SCZ GWAS (n = 254), using the PoPS method, were significantly enriched (fisher’s exact pvalue < 0.05, fisher’s OR > 1) in the GWAS risk genes from other approaches like nearest gene and MAGMA as well as burden-convergent genes for nearly all other disorders, except for PD (Figure S10A). In contrast, ASD genes prioritized from GWAS (n = 9) showed no significant enrichment in any burden-convergent gene sets (Figure S10B). We also found a strong and consistent enrichment between SCZ and BD convergence and known disease genes reflecting the strong genetic overlap observed between the disorders. Additionally, genes prioritized from both GWAS, and burden studies of AD were enriched only in convergent genes of AD and PD, implicating a distinct biological profile compared to the other psychiatric disorders (SCZ, ASD, & BD).

## Discussion

While GWAS and rare variant burden studies have independently linked common and rare variants to brain disorders, efforts to integrate these results to reveal shared biology remain limited. Here, we leverage convergent coexpression in post-mortem brain transcriptomes to uncover shared regulatory mechanisms underlying GWAS and rare variant burden risk across six neuropsychiatric diseases. Though GWAS and rare variant burden studies had limited overlap in their identified risk genes, we observed significant overlap of their convergent coexpressed genes. These convergent genes showed enrichment for both common and rare variant heritability. Moreover, genes convergently coexpressed with both GWAS and rare variant burden risk genes displayed greater evolutionary constraint and were more likely to be known drug targets compared to those exclusively convergent with either type. Notably, these shared convergent genes also pointed to disease-relevant cell types and biological processes.

The significant overlap between GWAS and rare variant burden convergent genes suggests that common and rare variants have shared downstream transcriptional effects. This is supported by prior evidence that convergent coexpression Z-scores correlate with gene expression changes from CRISPR perturbations^16^. Though differences in statistical power may lead GWAS and rare variant burden studies to prioritize different risk genes, convergent coexpression captures their complementary downstream effects. Notably, the degree of overlap between GWAS and rare burden convergent genes varied by disease, potentially reflecting differences in functional similarity and brain coexpression patterns of disease risk genes^49^. Our results highlight the tissue-specific nature of coexpression, indicating that common and rare variants may converge strongly in certain tissues or contexts. These context-dependent patterns could provide insights into how genetic variation contributes to disease in a more localized manner. Unlike module-based methods such as WGCNA^15^, that assess coexpression across the entire genome, our approach focuses on disease associated genes, enabling a targeted way to identify convergently coexpressed genes for further investigation.

Our findings suggest that GWAS and rare variant burden convergent genes could represent new risk genes, potentially reaching study-wide significance with increased sample size. Liao et al, had previously showed that convergence analyses for 71 burden genes included potential novel ASD genes that were not identified through current studies.^16^ Supporting this, we observed strong enrichment of both GWAS and rare burden convergent genes in both SNP-based and burden heritability, along with significant correlation with disease association p-values. Furthermore, GWAS-convergent genes identified using the PoPS approach were significantly enriched for known rare burden risk genes as well as GWAS risk genes identified through other approaches such as the nearest gene and MAGMA. Similarly, top burden-convergent genes for a given disorder showed enrichment for GWAS-prioritized genes. Collectively, these results emphasize the value of integrating convergence with common and rare variant studies to uncover risk genes.

Our approach also enabled the identification of shared biological processes between GWAS and rare variant burden studies. Limited number of genes reaching the exome-wide significance has posed a challenge to uncover biological mechanisms involved in rare variant burden studies. By leveraging convergent genes, we expanded the gene set available for more systematic investigation. The results revealed processes uniquely enriched in either GWAS or rare variant burden convergent genes, as well as those shared across both. Processes that showed enrichment in both types of convergent genes may represent core mechanisms and could point to systems involved in disease causality. We observed that across all six disorders, regulation of synaptic signal and vesicle release process showed enrichment in both GWAS and rare burden convergent genes underscoring established biology for the diseases. Enrichment of immune response and white blood cell proliferation processes supports previous observation of neuroinflammation as a central mechanism in AD pathogenesis^50,51^. Additionally, given prior evidence linking convergent coexpression to differential expression in cases vs controls, the potential of shared processes for patient stratification merits future investigation^16^.

Our analysis further revealed that genes converging with both GWAS, and rare variant burden risk genes tend to be more evolutionarily intolerant and were enriched for known drug targets, including those currently undergoing clinical investigation. This finding aligns with prior observations from Minikel et al, which showed drug targets supported by genetic evidence have a higher probability of success than those without such evidence^42^. The stronger enrichment observed in shared convergent genes also reinforces the findings from Sadler et al, which showed that integrating multiple layers of genetic approaches showed improved enrichment for drug target genes^50^. These findings suggest that disease-specific convergent genes and pathways supported by both common and rare variant evidence may enhance drug development strategies. Additionally, we observed that these shared convergent genes show high enrichment in cell-type marker gene sets, underscoring their functional relevance within specific cellular contexts and suggesting that targeting these genes could have cell-type-specific therapeutic effects.

Our study had several limitations. First, our analysis focuses on rare variant burden risk genes that have been discovered so far. As larger whole-exome sequencing studies are conducted, additional rare variant burden genes may be discovered, potentially revealing stronger overlaps with GWAS findings. Second, we relied on coexpression derived from bulk brain transcriptomic data from CMC and GTEx. Gene expression in bulk tissue can be obscured by cellular heterogeneity. Variant effects on transcriptomic data are highly context dependent such as cell type, cell states, and differentiation level^52^. Conducting convergence analysis using context specific coexpression may provide more precise insights into trait-relevant biological processes in higher resolutions. Additionally, our approach is also based on the previous observation that coexpression serves as a computational proxy for downstream effects of CRISPR perturbation outcomes. Nonetheless, we recognize that coexpression does not fully capture the complexity of perturbation outcomes, and experimental validation remains essential for identifying therapeutic targets. Finally, our analysis is limited to protein-coding genes harboring loss-of-function rare variants. It does not account for rare variants in noncoding regions, which play important regulatory role in gene expression.

In summary, we present a comprehensive approach to assess and quantify the overlap between GWAS and rare variant burden risk genes across six neuropsychiatric disorders. This approach is broadly generalizable to other complex traits beyond brain-related conditions and can help pinpoint genes influenced by both common and rare variant burden. By incorporating coexpression, our study highlights the importance of combining multiple layers of genetic evidence to uncover the functional convergence of risk genes and to deepen our understanding of disease mechanisms, which may ultimately implicate genes as potential therapeutic targets.

## Data Availability

All data produced in the present work are contained in the manuscript or available upon publication

## Acknowledgements

This work was funded by the National Institute of Mental Health (NIMH) and grant ID: R01MH123155.

## Supplementary figures

**Figure S1.**
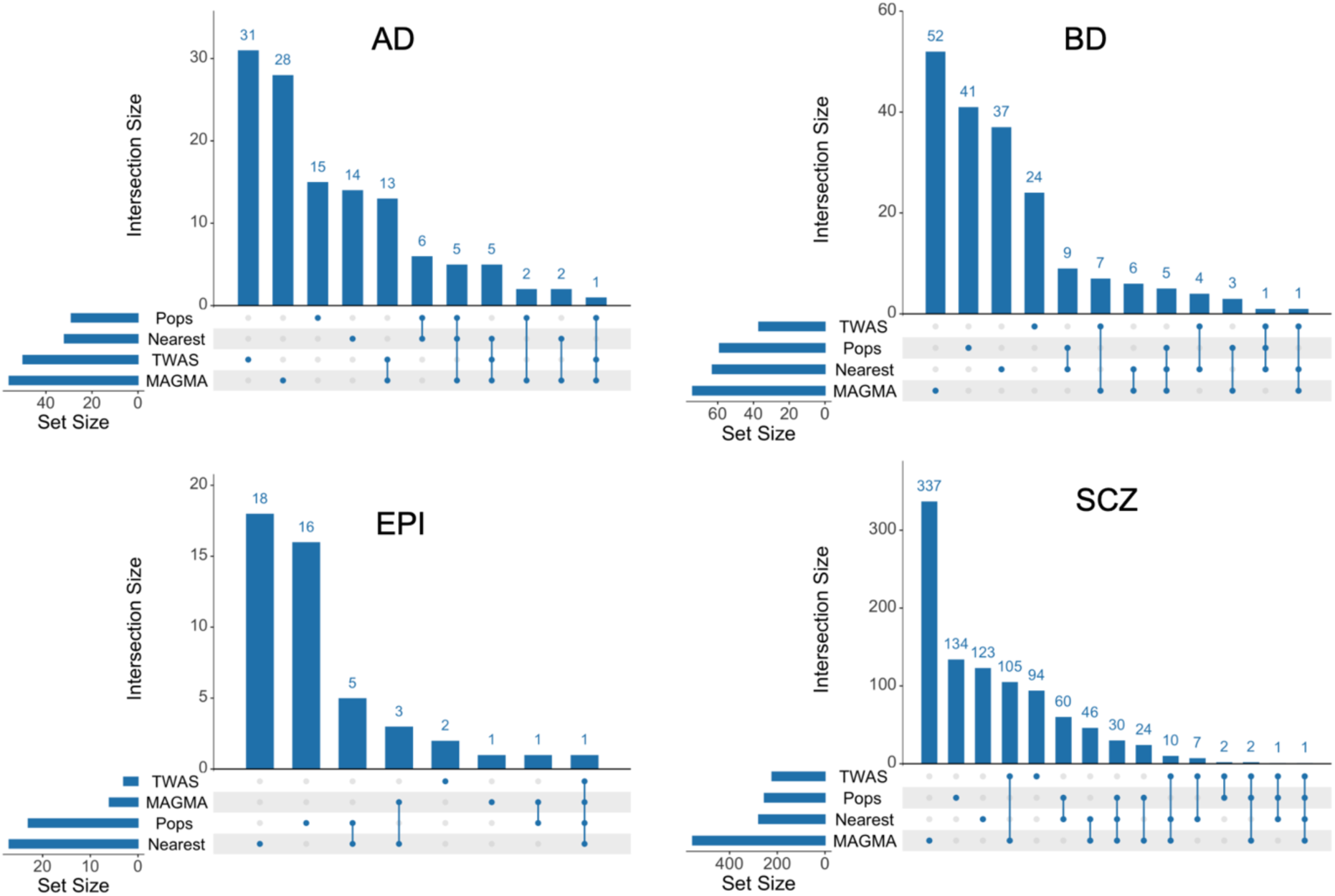
Upset plots showing overlap of prioritized GWAS genes across different approaches (TWAS, MAGMA, PoPS, and Nearest gene).

**Figure S2.**
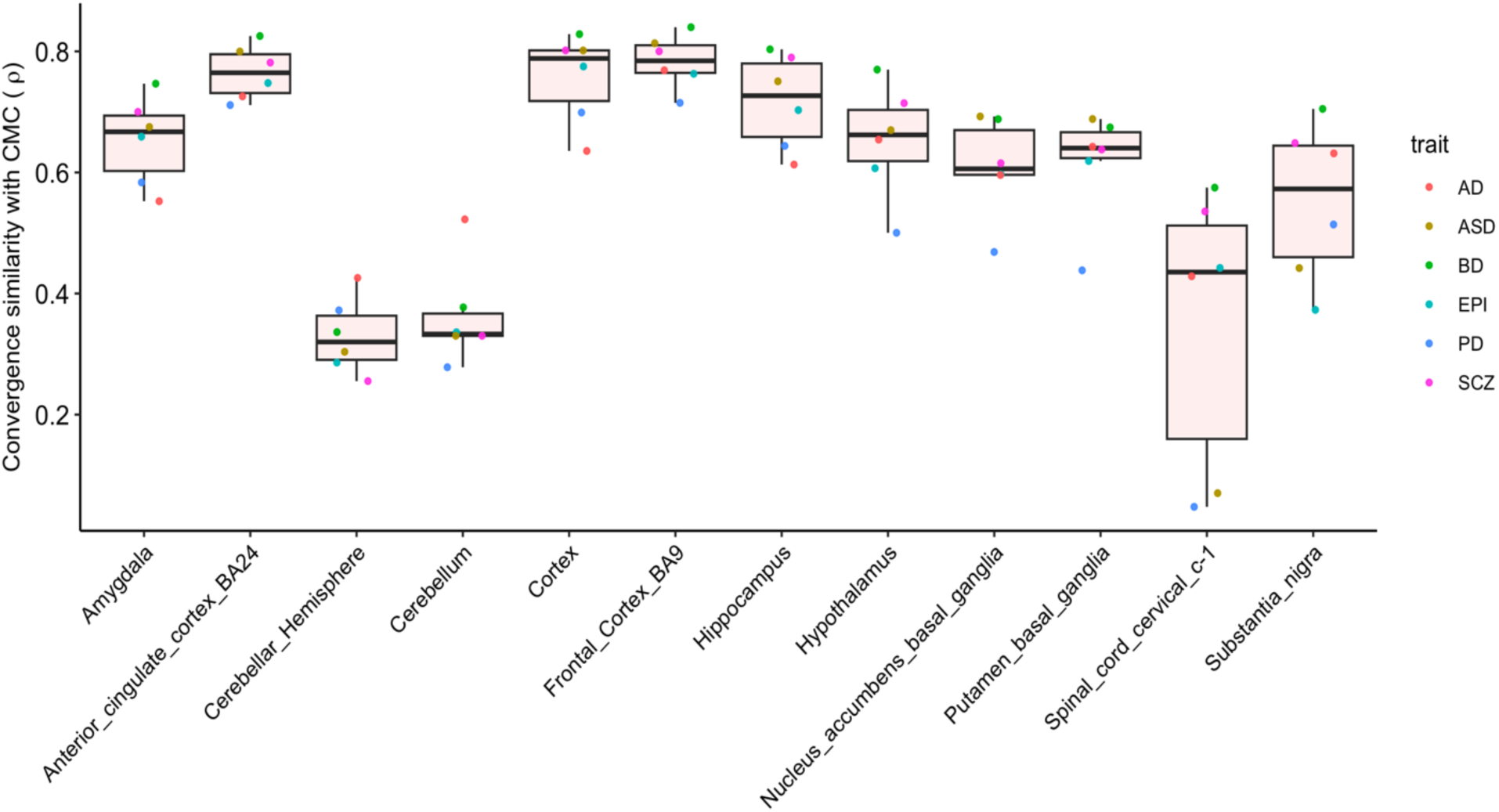
Similarity of burden convergence Z-score measured by spearman’s rho between that of CMC and 12 brain tissues from GTEx .

**Figure S3.**
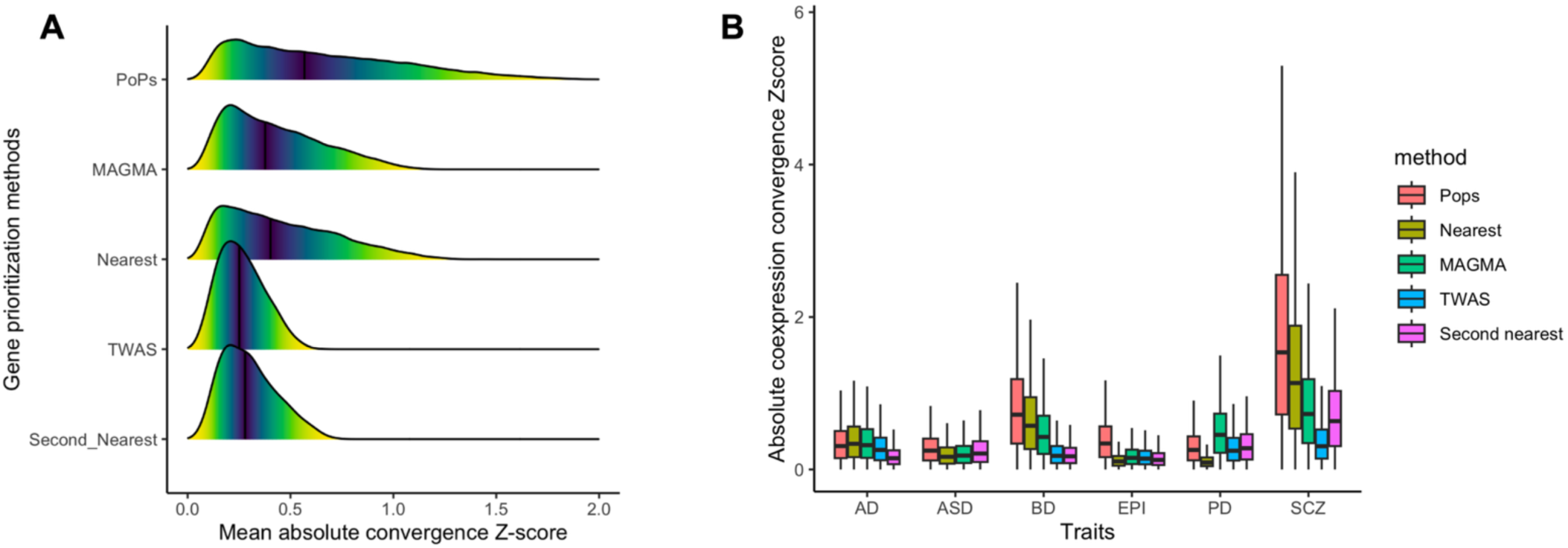
Comparison of convergence Z-score distributions across GWAS gene prioritization methods. A) Distribution of mean absolute convergence Z-scores across five prioritization methods. B) Distribution of mean absolute convergence Z-scores across five methods, stratified by disorder

**Figure S4.**
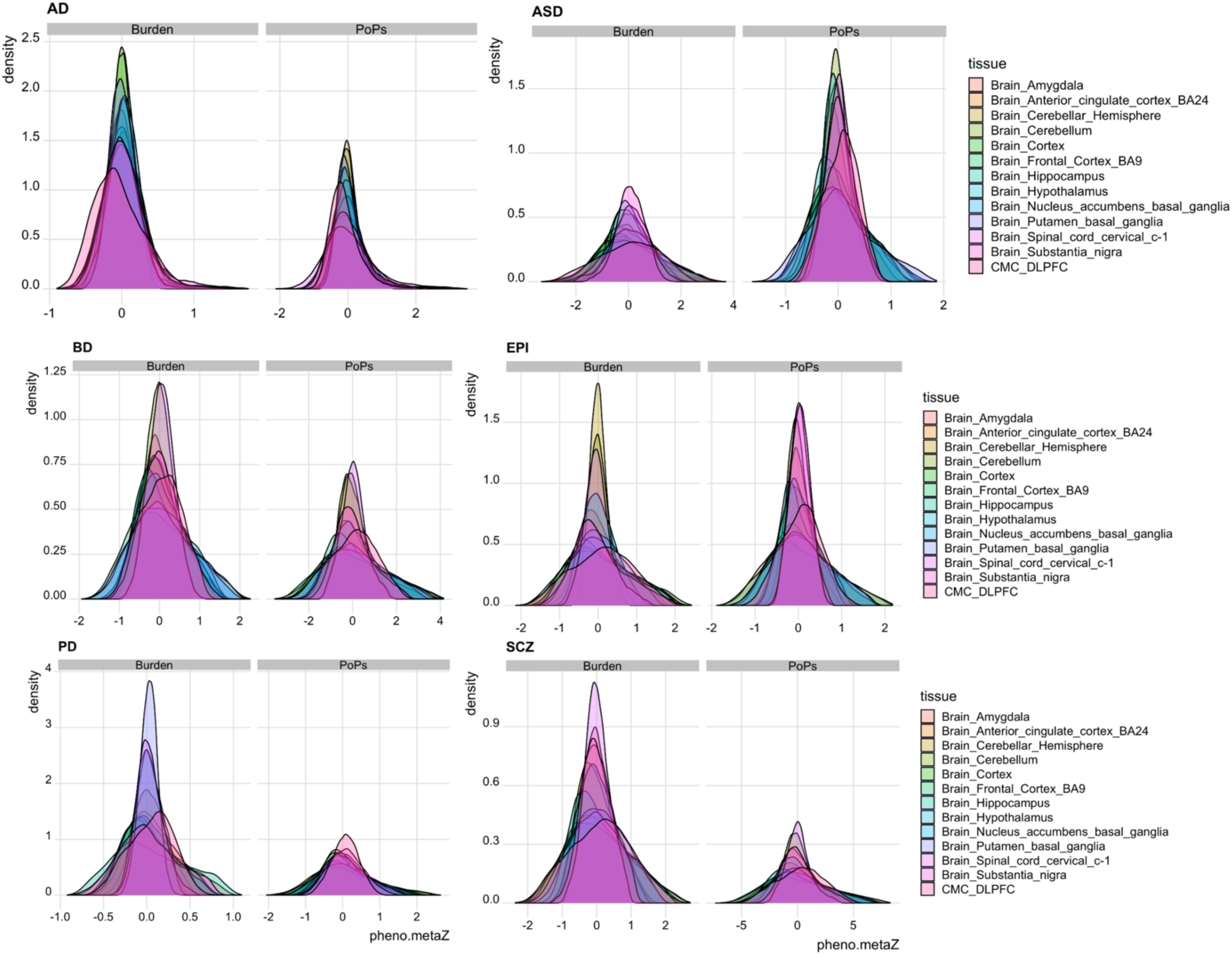
Distribution of GWAS (prioritized by PoPS) and rare variant burden convergent coexpression Zscores across 13 brain tissues from GTEx and CMC for the six brain disorders

**Figure S5.**
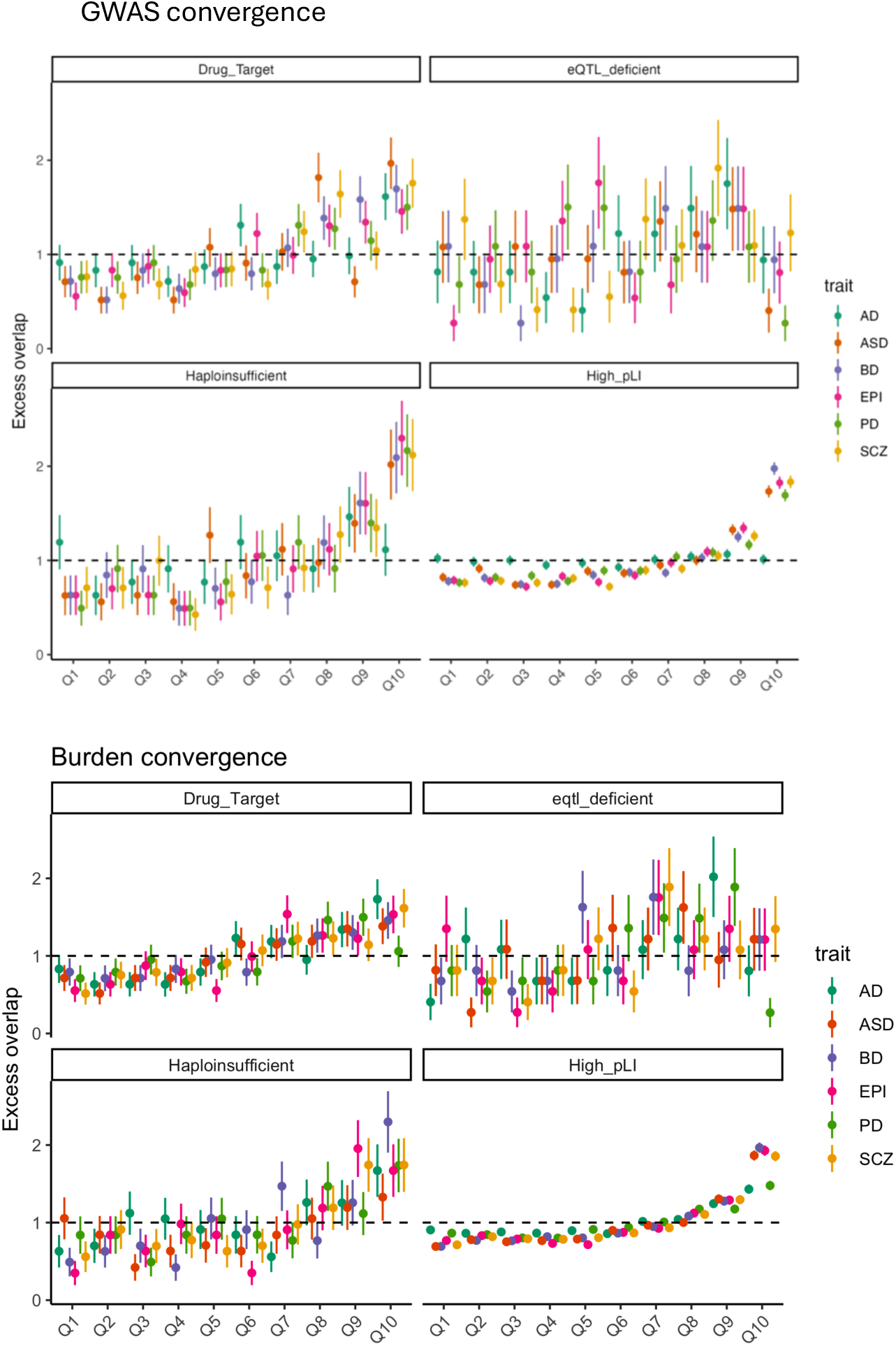
Excess overlap for essential disease genesets in both GWAS and rare variant burden convergence stratified by percentile of absolute convergence Zscore for the six disorders

**Figure S6.**
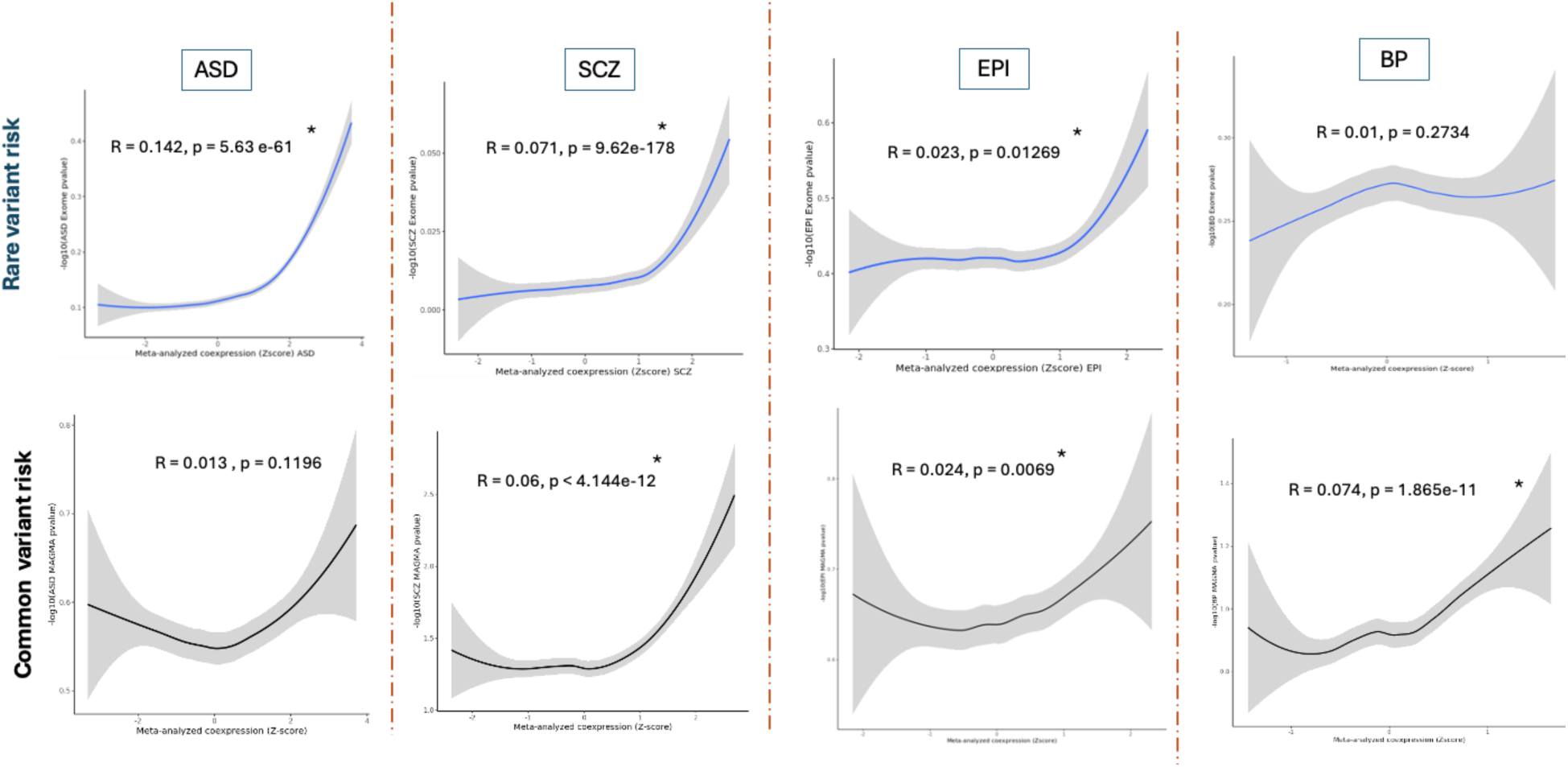
Correlation of burden convergence Zscores with exome and GWAS (MAGMA) association risk (pvalue)

**Figure S7.**
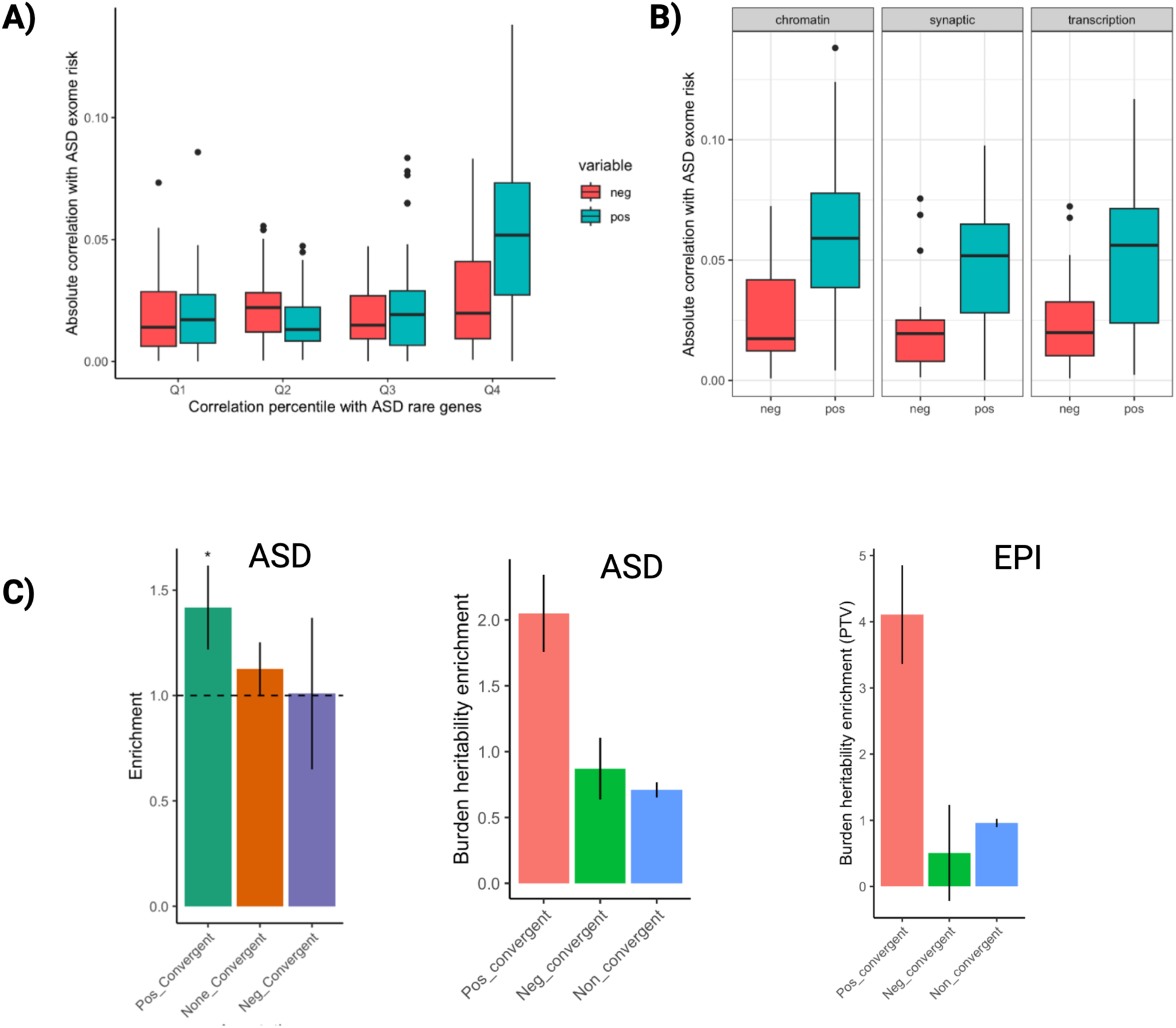
Stratification of Genes by Direction of Convergence Z-Scores A) Correlation between burden convergence Z-score and exome association risk (p-value) for genes stratified by Z-score direction in ASD. B) Correlation between burden convergence and exome p-value for genes grouped by functional categories and Z-score direction. C) Per-SNP and burden heritability of significantly convergent genes (p.bonf < 0.05) in ASD and EPI, categorized by Z-score direction.

**Figure S8.**
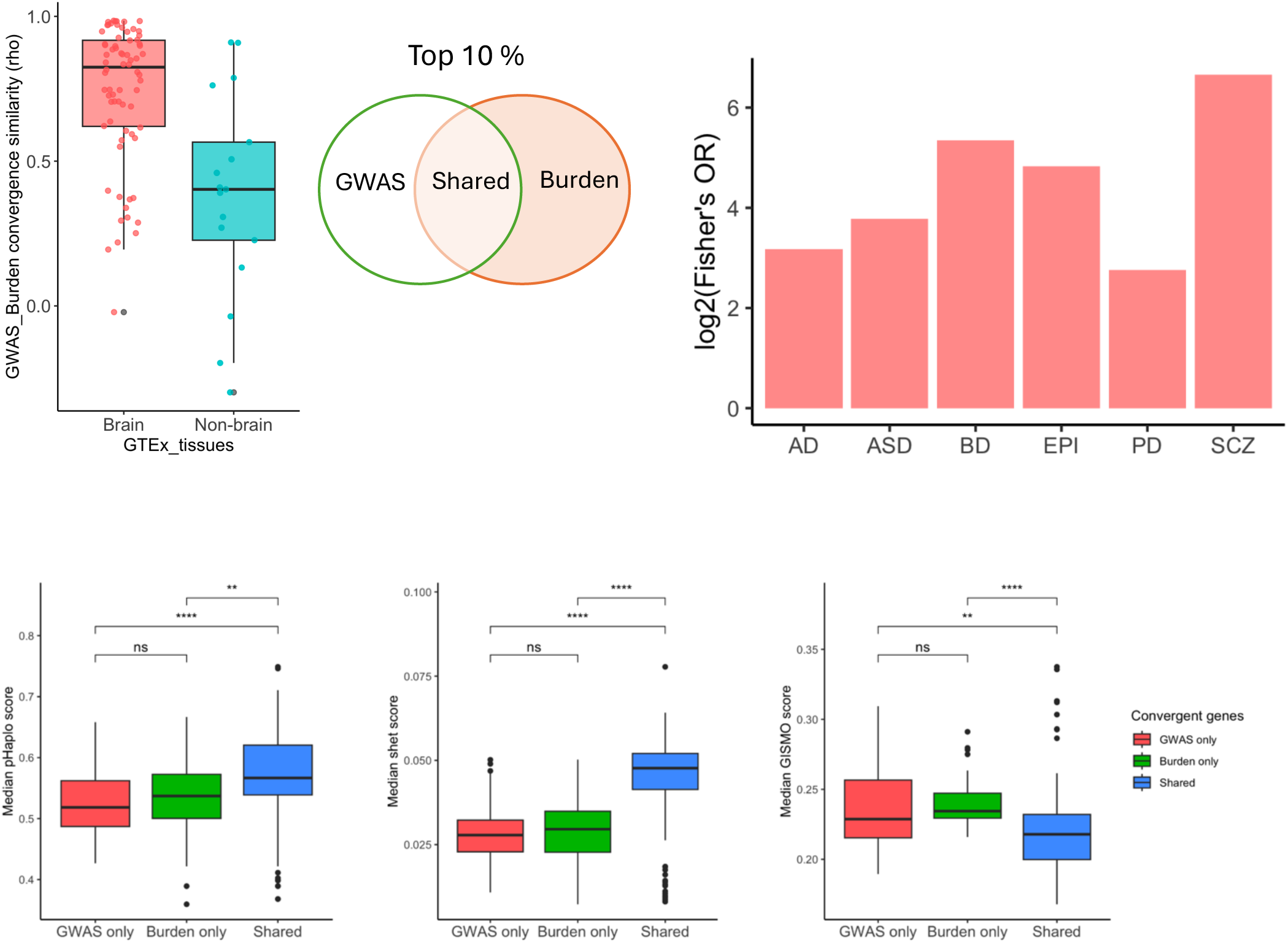
Median intolerance (constraint) scores, including LOUEF, pHaplo, shet, and GISMO calculated for the top decile of GWAS only, Burden only, and shared convergent genes across six neuropsychiatric diseases, using coexpression data from 13 brain tissues. Significance of distributions is calculated using Wilcox rank sum *(***p < 0.001, p < 0.01, p < 0.05, ns = not significant.)* For shet and pHaplo, the higher the score, the more intolerant; for GISMO and LOEUF, lower scores indicate intolerance

**Figure S9.**
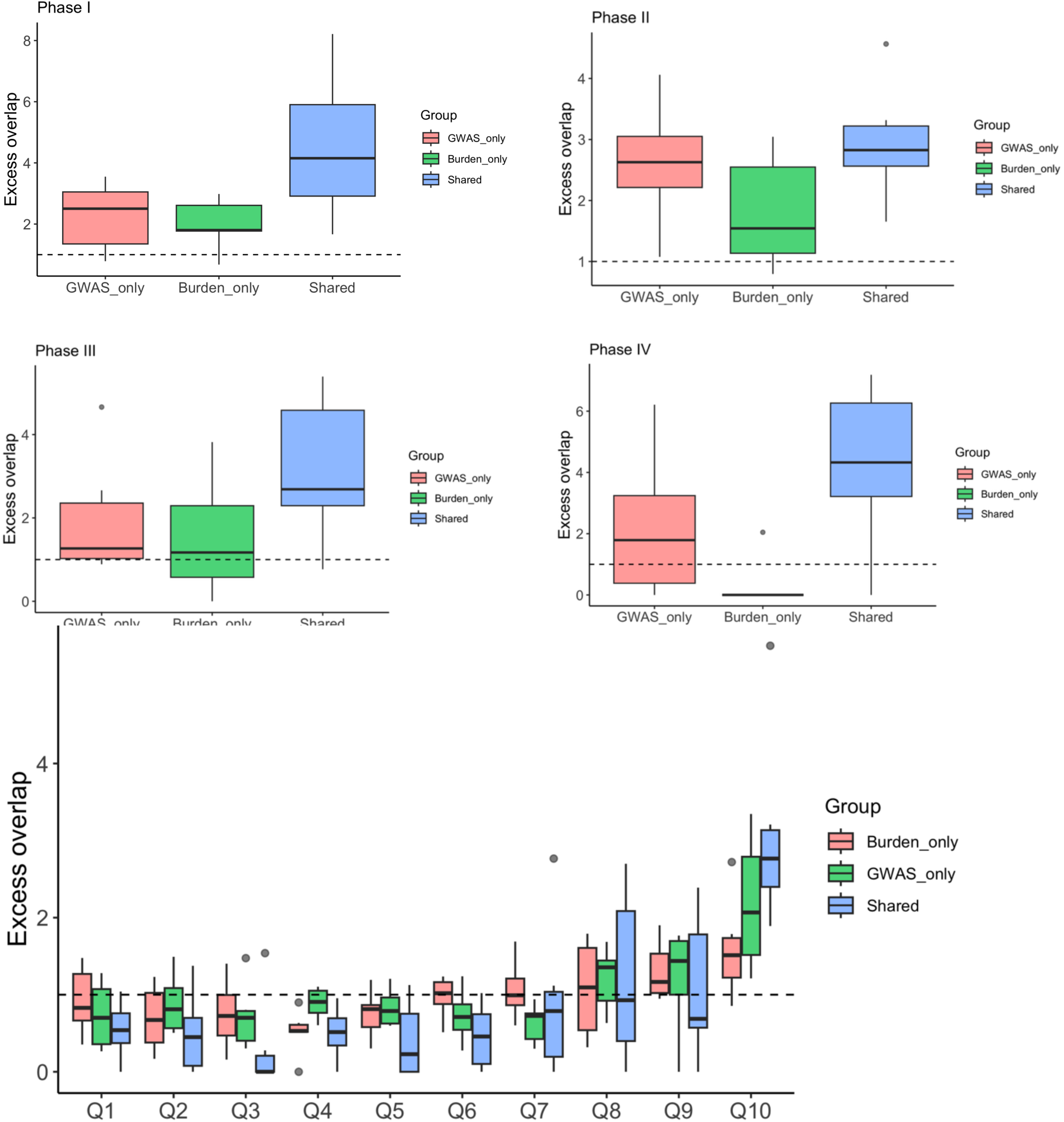
Excess overlap of known drug target genes at different clinical phases in convergent genes (A-D). Excess overlap of all drug target genes across the burden only, GWAS only, and Shared convergence divided by decile of absolute convergence Z-score

**Figure S10.**
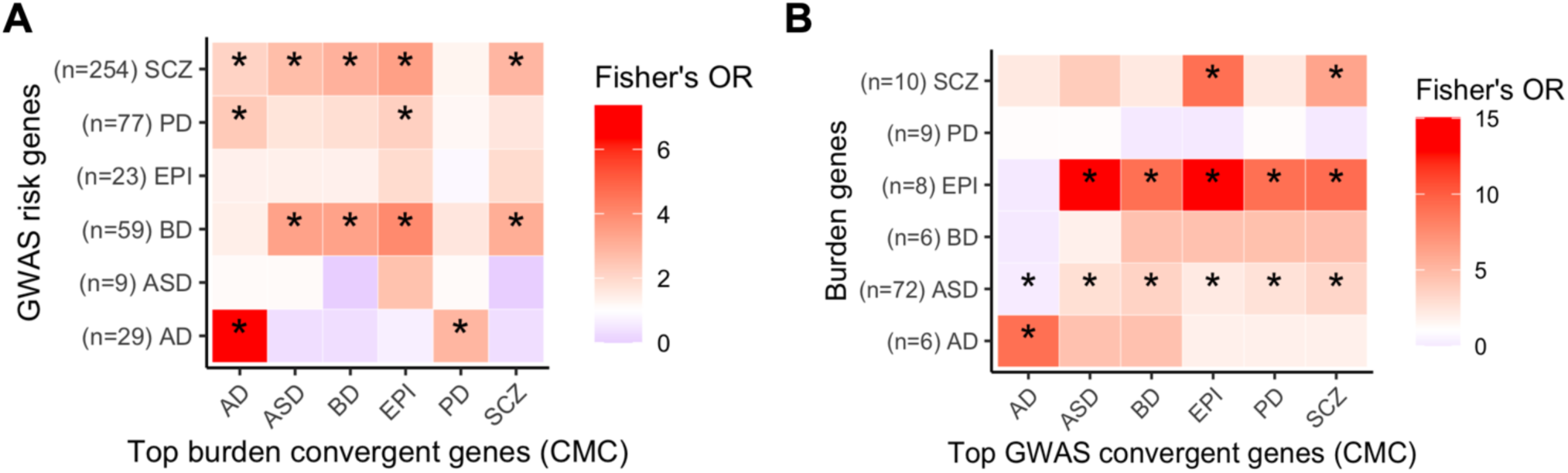
**A)** Enrichment of GWAS-prioritized risk genes among top rare variant burden convergent genes. The y-axis represents the number of prioritized GWAS risk genes. **B)** Enrichment of rare variant burden risk genes among top GWAS-convergent genes.

